# The prevalence, incidence and longevity of antibodies against SARS-CoV-2 among primary healthcare providers in Belgium: a prospective cohort study with 12 months of follow-up

**DOI:** 10.1101/2022.06.17.22276478

**Authors:** Niels Adriaenssens, Beatrice Scholtes, Robin Bruyndonckx, Pauline Van Ngoc, Jan Y Verbakel, An De Sutter, Stefan Heytens, Ann Van den Bruel, Isabelle Desombere, Pierre Van Damme, Herman Goossens, Laëtitia Buret, Els Duysburgh, Samuel Coenen

## Abstract

**Objectives:** To estimate the prevalence, incidence, and longevity of antibodies against SARS-CoV-2 among primary healthcare providers (PHCPs).

**Design:** Prospective cohort study with 12 months of follow-up.

**Setting:** Primary care in Belgium

**Participants:** Any general practitioner (GP) working in primary care in Belgium and any other PHCP from the same GP practice who physically manages (examines, tests, treats) patients were eligible. A convenience sample of 3,648 eligible PHCPs from 2,001 GP practices registered for this study (3,044 and 604 to start in December 2020 and January 2021, respectively). 3,390 PHCPs (92,9%) participated in their first testing timepoint (2,820 and 565, respectively) and 2,557 PHCPs (70,1%) in the last testing timepoint (December 2021).

**Interventions:** Participants were asked to perform a rapid serological test (RST) targeting IgM and IgG against the receptor binding domain (RBD) of SARS-CoV-2 and to complete an online questionnaire at each of maximum 8 testing timepoints.

**Primary and secondary outcome measures:** The prevalence, incidence, and longevity of antibodies against SARS-CoV-2 both after natural infection and after vaccination.

**Results:** Among all participants, 67% were women and 77% GPs. Median age was 43 years. The seroprevalence in December 2020 (before vaccination availability) was 15.1% (95% CI: 13.5% to 16.6%), increased to 84.2% (95% CI: 82.9% to 85.5%) in March 2021 (after vaccination availability) and reached 93.9% (95% CI: 92.9% to 94.9%) in December 2021 (during booster vaccination availability and fourth (delta variant dominant) covid wave). Among not (yet) vaccinated participants the first monthly incidence of antibodies against SARS-CoV-2 was estimated to be 2.91% (95% CI: 1.80% to 4.01%). The longevity of antibodies is higher in PHCPs with self-reported COVID-19 infection.

**Conclusions:** This study confirms that occupational health measures provided sufficient protection when managing patients. High uptake of vaccination resulted in high seroprevalence of SARS-CoV-2 antibodies in PHCPs in Belgium. Longevity of antibodies was supported by booster vaccination and virus circulation.

**Registration:** Trial registration number: NCT04779424

**Strengths and limitations of this study:**

- This large cohort study with 12 months follow-up could provide precise estimates of the prevalence and incidence of antibodies against SARS-CoV-2 among primary health care providers (PHCPs) at national and regional level in Belgium.
- The rapid serological test (RST) used targets IgM and IgG against the receptor binding domain of SARS-CoV-2 and could therefore also assess the antibody response after vaccination, and longevity of antibodies against SARS-CoV-2 both after natural infection and after vaccination, but cannot distinguish between both.
- The results in PHCPs could be compared to that of the general population and other population groups, e.g. health care workers in hospitals and nursing homes.
- The use of a convenience sample, missing data points and reduced RST accuracy when performed and interpreted by many different participants could limit the validity of the study results.

## Introduction

As of 8^th^ June 2022, severe acute respiratory syndrome coronavirus 2 (SARS-CoV-2) has caused over 530 million infections worldwide (4,164,698 in Belgium) and caused over 6.3 million deaths from coronavirus disease (COVID-19) worldwide (over 31,000 in Belgium).^1^ COVID-19 can be a lethal respiratory tract infection (RTI), but often presents with mild symptoms or remains asymptomatic.

Since the start of the COVID-19 pandemic, SARS-CoV-2 seroprevalence estimates have provided essential information about population exposure to infection and helped predict the early course of the epidemic.^2,3^ When setting up this study, seroprevalence studies in Iceland^4^ and Spain^5^ showed different levels of population antibody positivity, lasting up to at least 4 months in Iceland. In addition, early cohort studies have suggested waning of antibody levels in individuals is associated with, for example, illness severity, age and co-morbidities.^6-8^ Meanwhile, other seroprevalence studies showed antibody positivity lasting up to 9 months.^9,10^ Additionally, after vaccination, longevity of antibody positivity could differ depending on the type of vaccination and vaccination regime.^11,12^ For Belgium, Sciensano (the Belgian national institute of public health, www.sciensano.be) performs national seroprevalence studies of SARS-CoV-2 antibodies in the general population^13^ and several relevant populations including school-aged children and school staff,^14^ hospital staff,^15^ nursing homes residents and their staff.^16,17^ These results are publicly available and regularly updated on an online dashboard.^18^

This article focuses on the seroprevalence among primary healthcare providers (PHCPs).^19^ PHCPs manage the vast majority of patient contacts, including COVID-19 patients and therefore play an essential role in the efficient organisation of healthcare.^20,21^ Among the PHCPs, general practitioners (GPs) in particular, act as gatekeepers to the next levels of care. Therefore, preserving the capacity of GPs, together with that of their co-workers, throughout the COVID-19 epidemic is essential.^22^ In Belgium, this is particularly concerning given that the GP workforce consists of mainly older adults and is therefore at higher risk for COVID-19-related morbidity and mortality.^23^ In Italy, GPs represented up to 38% of the physicians who died from COVID-19 early in the epidemic.^24^

Before the start of this study (December 2020) data on how many PHCPs in Belgium had been infected by SARS-CoV-2 was not readily available,^25^ and effective vaccines for PHCPs were not anticipated to be available in the near future.

During the COVID-19 crisis rapid serological tests (RSTs) have been developed to identify the presence of antibodies to SARS-CoV-2. Compared to laboratory tests, a valid easy-to-use RST could speed up the availability of the test results for both the participants and the national health authorities.^25^ Furthermore, by using RSTs in this study, PHCPs got the opportunity to become more familiar with this type of technology.

Sciensano has validated five RSTs using finger prick blood, identifying one test with appropriate sensitivity (92.9%) and specificity (96.3%) for use in seroprevalence studies.^26^ We used this RST for the present study. It targets IgM and IgG against the receptor binding domain (RBD) of SARS-CoV-2 and could therefore also provide valuable information in a vaccinated population.

Given the availability of vaccines for PHCPs soon after the start of this study, we now report on the prevalence of antibodies against SARS-CoV-2 among a cohort of PHCPs in Belgium followed-up for 12-months, and on the incidence and longevity of those antibodies both after natural infection and after vaccination.

## Methods

This study was a prospective cohort study. Data collection was performed according to the publicly available protocol, providing more details on the study methods.^19^

### Study population

Any GP working in Belgium (including those in professional training) working in primary care and any PHCP from the same GP practice in a clinical role (clinical examination, testing or treating patients) were eligible if they were able to comply with the study protocol and provided informed consent to participate in the study. Staff hired on a temporary (interim) basis were excluded as follow-up over time would be compromised. Administrative staff or technical staff without any prolonged (longer than 15 minutes) face-to-face contact with patients and PHCPs who were not professionally active during the inclusion period were not eligible either.

PHCPs were recruited between 15 November 2020 and 15 January 2021. GPs working in clinical practice in Belgium were invited to register online for participation in this national epidemiological study and were asked to invite the other PHCPs in their practice to do the same. We emphasized that PHCPs who had already been diagnosed with COVID-19 were also eligible. Information about the study was disseminated to GPs and PHCPs via professional organisations (Domus Medica and Collège de Médecine Générale), university networks across the country and through professional media channels. We checked our convenience sample for representativeness in terms of geographic and demographic characteristics.^23^

To assess the geographical representativeness of our sample, we compared the distribution by region and by province of active GPs in Belgium in 2020 (source www.ima-aim.be) with the distribution of participating GPs.

### Data Collection

Upon inclusion in the study, participants were assigned a unique study code by the researchers and received testing material at their workplace through regular mail. At their first testing timepoint they received an invitation by email inviting them to auto-collect a capillary blood sample and analyse it using the RST (OrientGene®) and to complete a baseline questionnaire available in Dutch, French and English via a personalised link through a secured online platform hosted by Sciensano (Limesurvey). The invitation email included links to both written and video instructions to perform the RST on yourself and on someone else.

The baseline questionnaire at the first testing timepoint asked for informed consent and for information about the result of the RST, basic socio-demographic data (age, gender, composition of household – e.g. presence of school-aged children in the house), professional data (practice patient size), health status (pre-existing health conditions, regular medication use, presence of symptoms since the start of the epidemic, previous positive test results for COVID-19), professional exposure (contact with confirmed cases, use of infection prevention and control measures and the availability of personal protective equipment) and practice organisational aspects (delayed care for non-urgent conditions).^19^ A follow-up questionnaire was sent for each of the subsequent testing timepoints. In addition to the RST result, it collected information on the health status, including the presence of symptoms, COVID-19 testing and results, vaccination status (date of vaccination, type of vaccine, number of doses, presence of side-effects) and professional exposure (contact with confirmed cases, use of infection prevention and control measures).^19^

### Follow-up

The study lasted 12 months, from December 2020 to December 2021, and included 8 testing timepoints. Compared to the study protocol, the testing timepoint at the fifth month was skipped because of limited additional epidemiological value based on progressive insights from studies with similar protocols conducted by Sciensano that longer interval than four weeks between testing time point are suitable.^13-17^

### Sample size

This study aimed to include 5,000 PHCPs with a ratio of 4 GPs to 1 other PHCP. The sample size considerations regarding the different objectives of the proposed study are described in more detail in the study protocol.^19^ For the objectives reported here, even half the sample size aimed for would allow for precise estimates of the prevalence, incidence and longevity of antibodies against SARS-CoV-2.

### Data analysis

In the analysis, we included all PHCPs who provided informed consent and reported RST results at the testing timepoints. If in the questionnaire the entry for the date the RST was performed was missing or implausible, the date of completing the questionnaire was used instead. All analyses were conducted using R version 4.1.0 (www.R-project.org).

#### Prevalence

To assess the prevalence of antibodies against SARS-CoV-2, we calculated among the valid RST the proportion (95% CI) of positive RST for IgG and/or IgM, and for IgG and IgM separately (crude seroprevalences). In addition, we calculated the proportion (95% CI) of PHCPs that self-reported testing positive for SARS-CoV-2 (no test specified, so this includes both virus or antibody detection) since the outbreak of the COVID-19 pandemic (February 2020), and the proportion (95% CI) of PHCPs with any positive test, either a positive study RST or testing positive since the outbreak at their first testing timepoint. For any subsequent testing timepoints we asked the participants to specify if self-reported testing positive for SARS-CoV-2 since the previous testing timepoint concerned virus or antibody detection.

We also estimated the prevalence of antibodies against SARS-CoV-2 (IgG and/or IgM) taking into account clustering of PHCPs within their practice as well as the distribution of PHCPs across the districts in Belgium (adjusted seroprevalences). Weights were calculated based on the differences between the actual distribution of GPs across districts and the distribution of participating GPs with RST results across districts. These weights were then extrapolated to all other PHCPs. The estimates are based on Generalised Estimating Equations (GEE) assuming a binomial distribution for the RST result, an identity link function and an independent working correlation matrix.^27^ In a similar way we also estimated the adjusted prevalence of self-reported positive testing for SARS-CoV-2 since the start of the COVID-19 pandemic and the adjusted prevalence of these two tests results combined, either a positive study RST or testing positive since the outbreak for the first two testing timepoints.

#### Incidence

To assess the incidence of antibodies against SARS-CoV-2 (IgG and/or IgM) among participants not (yet) vaccinated, first we produced a Kaplan-Meier plot including participants providing a valid negative RST result at their first testing timepoint and not testing positive before, considering a positive RST during follow-up as event and censoring upon vaccination or loss to follow-up. Second, we assessed the monthly incidence of antibodies against SARS-CoV-2 due to natural infection in those not yet vaccinated, by analysing the data collected during the testing timepoints after the first testing timepoint. We included participants providing valid RST results both at the testing timepoint assessed and the preceding testing timepoint. We excluded participants reporting a positive RST at the preceding timepoint or already vaccinated with a first dose. In addition, we corrected the estimates for clustering of participants in general practices.

To assess the incidence of antibodies against SARS-CoV-2 (IgG and/or IgM) due to vaccination in those vaccinated, we calculated the proportion of participants with antibodies against SARS-CoV-2 less than seven days and seven days or more after the first, the second and the third dose of a COVID-19 vaccine, respectively, and stratified by self-reported history of COVID-19 infection.

#### Longevity

To assess the longevity of antibodies against SARS-CoV-2 (IgG and/or IgM) among participants not (yet) vaccinated, first we produced a Kaplan-Meier plot including participants without a self-reported history of COVID-19 infection before their first testing timepoint that provided a valid positive RST results before receiving their first dose of a COVID-19 vaccine, considering a negative RST result during follow-up as event (= negative RST result followed by another negative RST result or missing data) and censoring upon vaccination or loss to follow-up (midpoint and interval censoring). Second, we included participants not yet vaccinated, that provided a valid RST result at the testing timepoint assessed and a positive RST result at the previous testing timepoint. We estimated the proportion with a negative test result at the testing timepoint assessed.

To assess the longevity of antibodies against SARS-CoV-2 (IgG and/or IgM) after COVID-19 vaccination, we produced Kaplan-Meier plots by self-reported history of COVID-19 infection, including participants that provided a valid positive RST results at least seven days after receiving their second dose of a COVID-19 vaccine, considering a negative RST result during follow-up as event (= negative RST result followed by another negative RST result or missing data) and censoring upon booster vaccination (date of third dose) or loss to follow-up (midpoint and interval censoring).

### Vaccination

The start of the vaccination of PHCPs during the study follow-up provided the opportunity to monitor its progress.

### Ethics and dissemination

Ethical approval granted at 16 November 2020 (reference number: 20/46/605) by the Ethics Committee of the University Hospital Antwerp/University of Antwerp (Belgian registration number: 3002020000237).

During the study the information shown in Figure 1 was shared with the participants and the general population through the publicly available website of the Belgian health authorities (Sciensano) shortly after each testing-timepoint both for Belgium and its three regions, Brussels, Flanders and Wallonia.^18^

**Figure 1.**
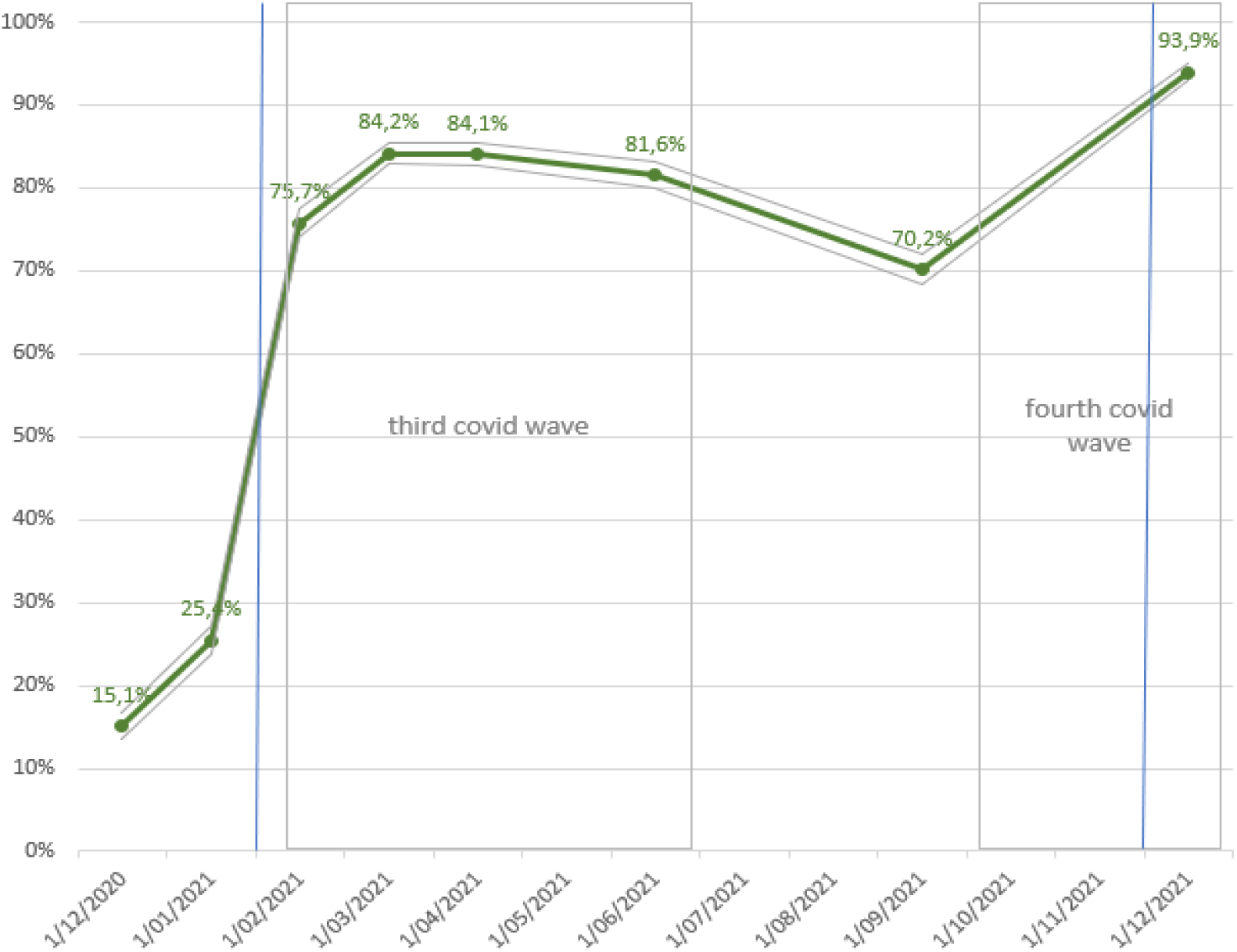
Prevalence of antibodies against SARS-CoV-2 among primary healthcare providers in Belgium from December 2020 to December 2021^1^ ^1^ The eight testing timepoints have the following start and end dates: T1: 24/12/2021-8/1/2021, T2: 25-31/1/2021, T3: 22-28/2/2021, T4: 22-31/3/2021, T5: 19-28/4/2021, T6: 14-27/6/2021, T7, 13-26/9/2021, T8: 13-26/12/2021. For the proportion of primary health care providers vaccinated at each testing timepoint see Table S4. The green line marks the prevalence of antibodies against SARS-CoV-2 (seroprevalence). The grey line mark the 95% confidence interval. The blue lines mark the start of primary and booster vaccination campaign for PHCPs. The grey boxes mark the third (15/2/2021-27/6/2021) and fourth COVID-19 (4/10/2021-27/12/2021).

## Results

### Description of the study cohort

In total, 3,648 eligible PHCPs from 2,001 practices registered and were asked to provide informed consent of whom 3,044 and 604 PHCPs were sent personal study materials to be able to collect data for their first testing timepoint starting on 24 December 2020 and 25 January 2021, respectively. 3,390 PHCPs participated in their first testing timepoint by completing the baseline questionnaire, among which 2,597 GPs, 386 GPs in training and 407 other PHCPs (Table 1).

**Table 1.** Characteristics of primary healthcare providers (PHCPs), including general practitioners (GPs), GPs in training and other PHCPs who participated in their first testing timepoints^1^.

|  | PHCPs<br>n=3,390 |  | GPs<br>n=2,597 |  | GPs in training<br>n=386 |  | Other PHCPs<br>n=407 |  |
| --- | --- | --- | --- | --- | --- | --- | --- | --- |
| Age<sup>2</sup>, median (IQR) | 40 | (31-54) | 44 | (34-57) | 27 | (26-28) | 38 | (31-47) |
| Gender<sup>3</sup>, n (%) |  |  |  |  |  |  |  |  |
| - Male | 1,119 | (33.0) | 943 | (36.3) | 112 | (29.0) | 64 | (15.7) |
| - Female | 2,296 | (66.9) | 1,652 | (63.6) | 274 | (71.0) | 343 | (84.3) |
| - Not reported | 2 | (0.1) | 2 | (0.1) | 0 | (0) | 0 | (0) |
| Practice size, n (%)<sup>3</sup> |  |  |  |  |  |  |  |  |
| - Solo | 618 | (33.5) | 580 | (34.7) | 54 | (16.1) | 29 | (11.8) |
| - Duo | 361 | (19.6) | 328 | (19.6) | 74 | (22.1) | 32 | (13.1) |
| - Group (<8 employees) | 382 | (20.7) | 351 | (21.0) | 51 | (15.2) | 21 | (8.6) |
| - Large group (>7 employees) | 444 | (24.1) | 386 | (23.1) | 156 | (46.6) | 150 | (61.2) |

<sup>1</sup> The first testing timepoint was December 2020 for 2,820 and January 2021 for 570 PHCPs, respectively; <sup>2</sup>Ages < 21 were considered unrealistic and recoded as missing; IQR=interquartile range; <sup>3</sup> if numbers do not add up to the column total, this is due to missing data; numbers of practices for PHCPs=1,845, GPs=1,672, GPs in training=335 and other PHCPs=245.

Our sampling procedure resulted in the participation of a reasonably geographically representative sample of GPs at the level of the provinces (Table S1, online supplementary data). At the level of the regions, there is about 8% overrepresentation of GPs in Flanders and corresponding underrepresentation of GPs in Wallonia.

### Participant characteristics

Table 1 presents the characteristics of the 3,390 PHCPs who participated in their first (baseline) testing timepoint. These PHCPs, mainly GPs, were relatively young, more often female and working more often in (large) group practices than in solo or duo practices. Table 2 shows in how many testing timepoints primary healthcare providers (PHCPs) participated. 3,415 (93.6%) PHCPs participated in at least one testing timepoint, 2,909 (79,7%) participated in six and 2,141 (58.7%) participated in all eight testing timepoints. The number of PHCPs participating per testing timepoint is presented in Table S2 (online supplementary data). While the response rate gradually decreased, still 2,557 (77.2% of invited PHCPs) participated in the last testing timepoint.

**Table 2.** The number of testing timepoints that primary healthcare providers (PHCPs) participated in.

| Number of testing timepoints participated in | Number of PHCPs (%)<br>N=3,648 |  | Cumulative percentage |
| --- | --- | --- | --- |
| 8<sup>1</sup> | 2,141 | (58.7%) | 58.7% |
| 7 | 490 | (13.4%) | 72.1% |
| 6 | 278 | (7.6%) | 79.7% |
| 5 | 153 | (4.2%) | 83.9% |
| 4 | 129 | (3.5%) | 87.5% |
| 3 | 91 | (2.5%) | 90.0% |
| 2 | 87 | (2.4%) | 92.4% |
| 1 | 46 | (1.3%) | 93.6% |
| 0 | 233 | (6.4%) | 100.0% |

<sup>1</sup> The eight testing timepoints have the following start and end dates: T1: 24/12/2021-8/1/2021, T2: 25-31/1/2021, T3: 22-28/2/2021, T4: 22-31/3/2021, T5: 19-28/4/2021, T6: 14-27/6/2021, T7, 13-26/9/2021, T8: 13-26/12/2021.

### Vaccination status

Overall, 3,227 participants received a full primary vaccination. 2,783 participants received two doses of an m-RNA vaccine (2,639 (81.8%) BNT162b2, 144 (4.5%) mRNA-1273 and 2 (0.1%) mRNA-1273 followed by BNT162b2). 437 participants (13.5%) received two doses of ChAdOx1-S and 5 (0.2%) participants one dose of Ad26.COV2.S.

At the final testing timepoint, 2,211 of the participants had received a booster vaccination. 1,879 (85.0%) participants received a booster with BNT162b2 and 267 (12.1%) with mRNA-1273. 1 participant received ChAdOx1-S and another participant Ad26.COV2.S as third dose.

**Table S1.** Distribution by province of active general practitioners (GPs) in Belgium in 2020 and of GPs who participated in CHARMING in their testing timepoint^1^.

| Region/Province | Active GPs<br>n (%) |  | Participating GPs<br>n (%) |  |
| --- | --- | --- | --- | --- |
| Brussels | 1,178 | (10.01) | 239 | (9.2) |
| Flanders | 6,805 | (57.83) | 1,725 | (66.4) |
| Wallonia | 3,784 | (32.16) | 633 | (24.4) |
| Antwerpen-Anvers | 1,806 | (15.35) | 454 | (17.5) |
| Brussel-Hoofdstad-Bruxelles Capitale | 1,178 | (10.01) | 239 | (9.2) |
| Henegouwen-Hainaut | 1,293 | (10.99) | 175 | (6.7) |
| Limburg-Limbourg | 943 | (8.01) | 235 | (9.0) |
| Luik-Liège | 1,125 | (9.56) | 200 | (7.7) |
| Luxemburg-Luxembourg | 301 | (2.56) | 78 | (3.0) |
| Namen-Namur | 594 | (5.05) | 104 | (4.0) |
| Oost-Vlaanderen-Flandre Orientale | 1,556 | (13.22) | 431 | (16.6) |
| Vlaams-Brabant-Brabant-Flamand | 1,241 | (10.55) | 317 | (12.2) |
| Waals-Brabant-Brabant Wallon | 471 | (4.00) | 76 | (2.9) |
| West-Vlaanderen-Flandre Occidentale | 1,259 | (10.70) | 288 | (11.1) |
| Total | 11,767 |  | 2,597 | (22.1) |

<sup>†</sup> The first testing timepoint was December 2020 for 2224 and January 2021 for 373 GPs. PHCPs, respectively.

**Table S2.**
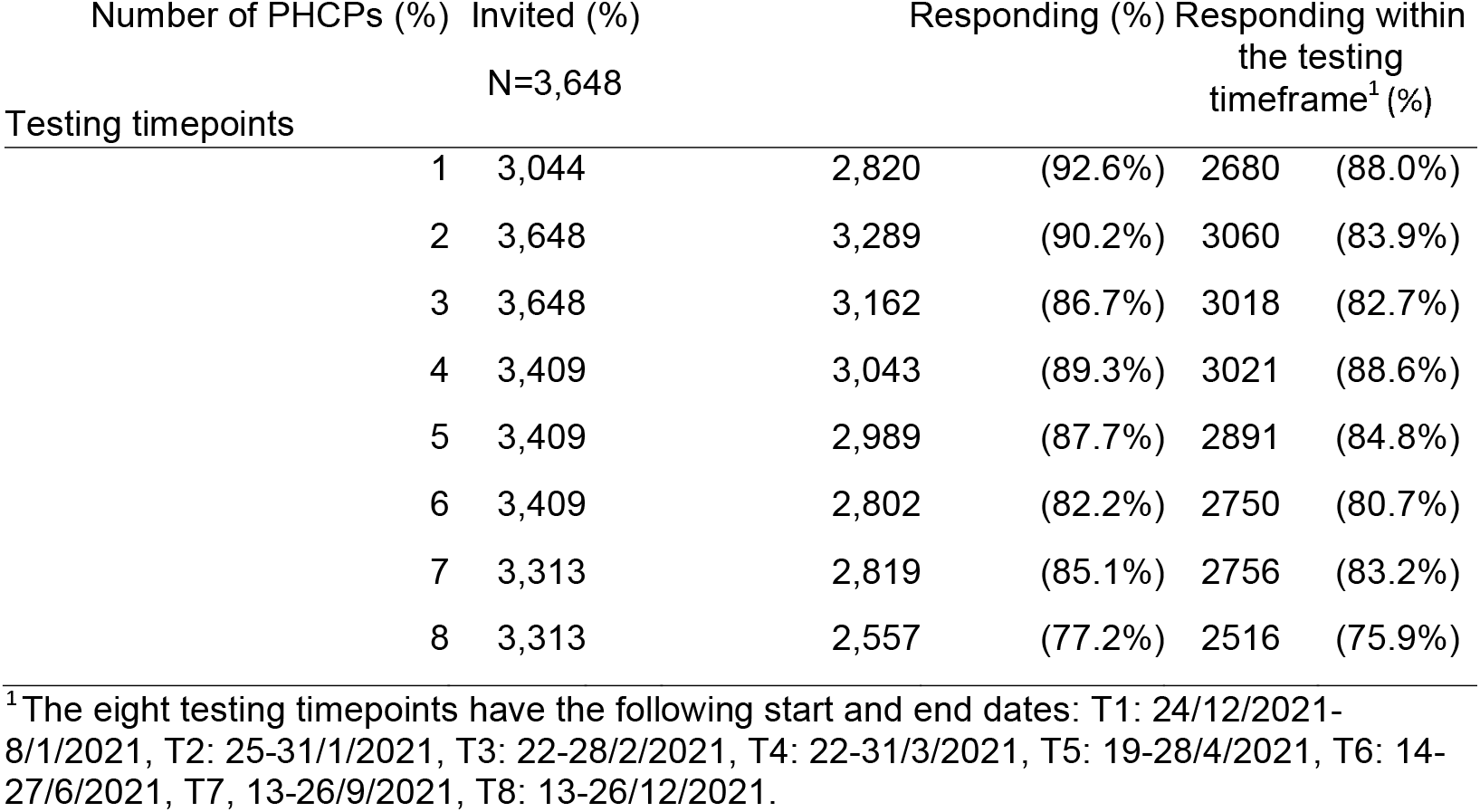
The number of primary healthcare providers (PHCPs) participating per testing timepoint.

### Prevalence

The prevalence of antibodies against SARS-CoV-2 among PHCPs in Belgium from December 2020 to December 2021 is shown in Figure 1 and Table S3. Table S3 also gives the number of eligible PHCPs, i.e. those testing between the start and end date of the respective testing timepoint, as well as the regional differences. At the first testing timepoint (T1), among 2680 eligible PHCPs, 2629 provided valid test results, of which 366 (15.1%) were positive. Afterwards, the prevalence increased substantially up to 84.2% at T4, mainly due to vaccination (see Table S4). Six months later (T7) the prevalence was substantially lower (70.2%), while during the fourth covid wave (delta variant dominant) and after booster vaccination became available it increased again to 93.9% (T8).

**Table S3.** Prevalence of antibodies against SARS-CoV-2 among primary healthcare providers in Belgium at eight testing timepoints from December 2020 to December 2021^1^.

| Testing timepoint | Region | PHCPs n | Valid RST<sup>2</sup> n | Positive RST<sup>3</sup> n | Adjusted prevalence<sup>4</sup> % (95% CI) |  |
| --- | --- | --- | --- | --- | --- | --- |
| T1 | Belgium | 2,680 | 2,629 | 366 | 15.08 | (13.54-16.62) |
|  | Brussels | 234 | 233 | 43 | 18.45 | (13.47-23.44) |
|  | Flanders | 1841 | 1800 | 203 | 11.28 | (9.77-12.79) |
|  | Wallonia | 605 | 596 | 120 | 20.37 | (16.91-23.84) |
| T2 | Belgium | 3,060 | 2,995 | 716 | 25.42 | (23.75-27.08) |
|  | Brussels | 270 | 263 | 55 | 20.91 | (15.98-25.84) |
|  | Flanders | 2024 | 1980 | 389 | 20.10 | (18.29-21.92) |
|  | Wallonia | 766 | 752 | 272 | 36.03 | (32.46-39.60) |
| T3 | Belgium | 3,018 | 2,967 | 2,278 | 75.70 | (74.03-77.37) |
|  | Brussels | 274 | 273 | 168 | 61.54 | (55.72-67.36) |
|  | Flanders | 2014 | 1971 | 1615 | 82.35 | (80.63-84.06) |
|  | Wallonia | 730 | 723 | 495 | 68.80 | (65.26-72.34) |
| T4 | Belgium | 3,021 | 2,980 | 2,509 | 84.17 | (82.86-85.48) |
|  | Brussels | 279 | 274 | 209 | 76.28 | (71.24-81.31) |
|  | Flanders | 1,987 | 1,963 | 1,706 | 86.91 | (85.42-88.40) |
|  | Wallonia | 755 | 743 | 594 | 79.95 | (77.07-82.83) |
| T5 | Belgium | 2,891 | 2,859 | 2,410 | 84.07 | (82.65-85.48) |
|  | Brussels | 274 | 268 | 206 | 76.87 | (71.82-81.91) |
|  | Flanders | 1,898 | 1,877 | 1,622 | 86.67 | (85.09-88.25) |
|  | Wallonia | 719 | 714 | 582 | 81.86 | (78.97-84.75) |
| T6 | Belgium | 2,750 | 2,725 | 2,230 | 81.57 | (80.02-83.12) |
|  | Brussels | 252 | 244 | 197 | 80.74 | (75.81-85.67) |
|  | Flanders | 1,839 | 1,826 | 1,514 | 82.76 | (80.97-84.55) |
|  | Wallonia | 659 | 655 | 519 | 79.78 | (76.59-82.98) |
| T7 | Belgium | 2,756 | 2,730 | 1,917 | 70.17 | (68.36-71.97) |
|  | Brussels | 238 | 237 | 178 | 75.11 | (69.62-80.59) |
|  | Flanders | 1,844 | 1,823 | 1,271 | 69.38 | (67.20-71.56) |
|  | Wallonia | 674 | 670 | 468 | 70.04 | (66.46-73.62) |
| T8 | Belgium | 2,516 | 2,498 | 2,356 | 93.91 | (92.89-94.93) |
|  | Brussels | 222 | 221 | 201 | 90.95 | (87.17-94.73) |
|  | Flanders | 1,696 | 1,681 | 1,607 | 95.42 | (94.36-96.47) |
|  | Wallonia | 598 | 596 | 548 | 92.22 | (90.02-94.43) |

<sup>1</sup> See Table S4 for the proportions of PHCPs partially and fully vaccinated; <sup>2</sup> RST: Rapid Serological Test; <sup>3</sup> IgG and/or IgM positive among the valid RST; <sup>4</sup> Estimates are based on Generalised Estimating Equations taking into account clustering of PHCPs within their practice and distribution of GPs across districts in Belgium; T1: 24/12/2021-8/1/2021, T2: 25-31/1/2021, T3: 22-28/2/2021, T4: 22-31/3/2021, T5: 19-28/4/2021, T6: 14-27/6/2021, T7, 13-26/9/2021, T8: 13-26/12/2021.

**Table S4.** Proportions of primary healthcare providers in Belgium with valid rapid serological test results^1^ vaccinated at eight testing timepoints from December 2020 to December 2021.

| Testing timepoint | Region | Partially vaccinated<sup>2</sup><br>% (95%CI) |  | Fully vaccinated<sup>3</sup><br>% (95%CI) |  | Booster vaccinated<sup>4</sup><br>% (95%CI) |  |
| --- | --- | --- | --- | --- | --- | --- | --- |
| T1 | Belgium | NA |  | NA |  | NA |  |
|  | Brussels | NA |  | NA |  | NA |  |
|  | Flanders | NA |  | NA |  | NA |  |
|  | Wallonia | NA |  | NA |  | NA |  |
| T2 | Belgium | 57.16 | (55.39-58.93) | 0.87 | (0.54-1.20) | NA |  |
|  | Brussels | 17.49 | (12.90-22.08) | 1.14 | (0.00-2.42) | NA |  |
|  | Flanders | 67.27 | (65.21-69.34) | 0.30 | (0.06-0.55) | NA |  |
|  | Wallonia | 44.41 | (40.86-47.97) | 2.26 | (1.20-3.32) | NA |  |
| T3 | Belgium | 16.92 | (15.57-18.27) | 66.23 | (64.53-67.93) | NA |  |
|  | Brussels | 50.18 | (44.25-56.11) | 21.98 | (17.07-26.89) | NA |  |
|  | Flanders | 11.72 | (10.30-13.14) | 76.15 | (74.27-78.04) | NA |  |
|  | Wallonia | 18.53 | (15.70-21.37) | 55.88 | (52.26-59.50) | NA |  |
| T4 | Belgium | 16.88 | (15.53-18.22) | 78.46 | (76.98-79.93) | NA |  |
|  | Brussels | 30.29 | (24.85-35.73) | 60.22 | (54.42-66.01) | NA |  |
|  | Flanders | 13.40 | (11.89-14.90) | 84.06 | (82.44-85.67) | NA |  |
|  | Wallonia | 21.13 | (18.20-24.07) | 70.39 | (67.11-73.67) | NA |  |
| T5 | Belgium | 15.49 | (14.17-16.82) | 81.11 | (79.68-82.55) | NA |  |
|  | Brussels | 26.46 | (22.21-31.78) | 66.04 | (60.38-71.71) | NA |  |
|  | Flanders | 13.16 | (11.63-14.69) | 85.35 | (83.75-86.95) | NA |  |
|  | Wallonia | 17.51 | (14.72-20.29) | 80.48 | (77.13-83.83) | NA |  |
| T6 | Belgium | 1.54 | (1.08-3.00) | 95.93 | (95.18-96.67) | NA |  |
|  | Brussels | 2.05 | (0.27-3.83) | 92.21 | (88.85-95.58) | NA |  |
|  | Flanders | 0.82 | (0.41-1.24) | 98.68 | (97.39-98.67) | NA |  |
|  | Wallonia | 3.36 | (1.42-5.35) | 92.90 | (90.63-95.17) | NA |  |
| T7 | Belgium | 0.51 | (0.24-0.78) | 97.91 | (97.38-98.45) | 0.73 | (0.41-1.05) |
|  | Brussels | 2.53 | (0.53-4.53) | 94.51 | (91.62-97.41) | 0.42 | (0.00-1.25) |
|  | Flanders | 0.11 | (0.00-0.26) | 99.23 | (98.88-99.63) | 0.93 | (0.49-1.37) |
|  | Wallonia | 0.90 | (0.18-1.61) | 95.52 | (93.96-97.09) | 0.30 | (0.00-0.71) |
| T8 | Belgium | 0.20 | (0.02-0.38) | 98.72 | (98.28-99.16) | 84.78 | (83.37-86.18) |
|  | Brussels | 0.00 | (0.00-0.00) | 98.64 | (97.12-100.00) | 72.07 | (66.17-77.97) |
|  | Flanders | 0.06 | (0.00-0.18) | 99.46 | (99.12-99.81) | 89.74 | (88.30-91.18) |
|  | Wallonia | 0.67 | (0.02-1.33) | 96.64 | (95.20-98.09) | 75.42 | (71.97-78.87) |

<sup>1</sup> See Table S3 for the number of primary healthcare providers with valid rapid serological test results; <sup>2</sup> Received one out of two doses; <sup>3</sup> Received two doses; <sup>4</sup> Received a third dose. T1: 24/12/2021-8/1/2021, T2: 25-31/1/2021, T3: 22-28/2/2021, T4: 22-31/3/2021, T5: 19-28/4/2021, T6: 14-27/6/2021, T7, 13-26/9/2021, T8: 13-26/12/2021.

### Incidence

#### Among not (yet) vaccinated participants

The incidence of antibodies against SARS-CoV-2 among PHCPs in Belgium among participants that provided a valid negative RST result at their first testing timepoint, did not self-report a COVID-19 infection before and were not (yet) vaccinated is shown in figure 2. For the second testing timepoint (T2) the monthly incidence of antibodies against SARS-CoV-2 was estimated to be 2.91% (95%CI: 1.80-4.01; n=895), i.e. the proportion of PHCPs not yet vaccinated at T2 and testing negative at T1, that tested positive at T2. For T3 and T4 it was estimated to be 3.93% (95%CI: 2.04-5.82; n=407) and 4.04% (95%CI: 0.16 - 7.92; n=99), respectively. As of T4, the sample size of eligible participants was too small for precise estimates.

**Figure 2.**
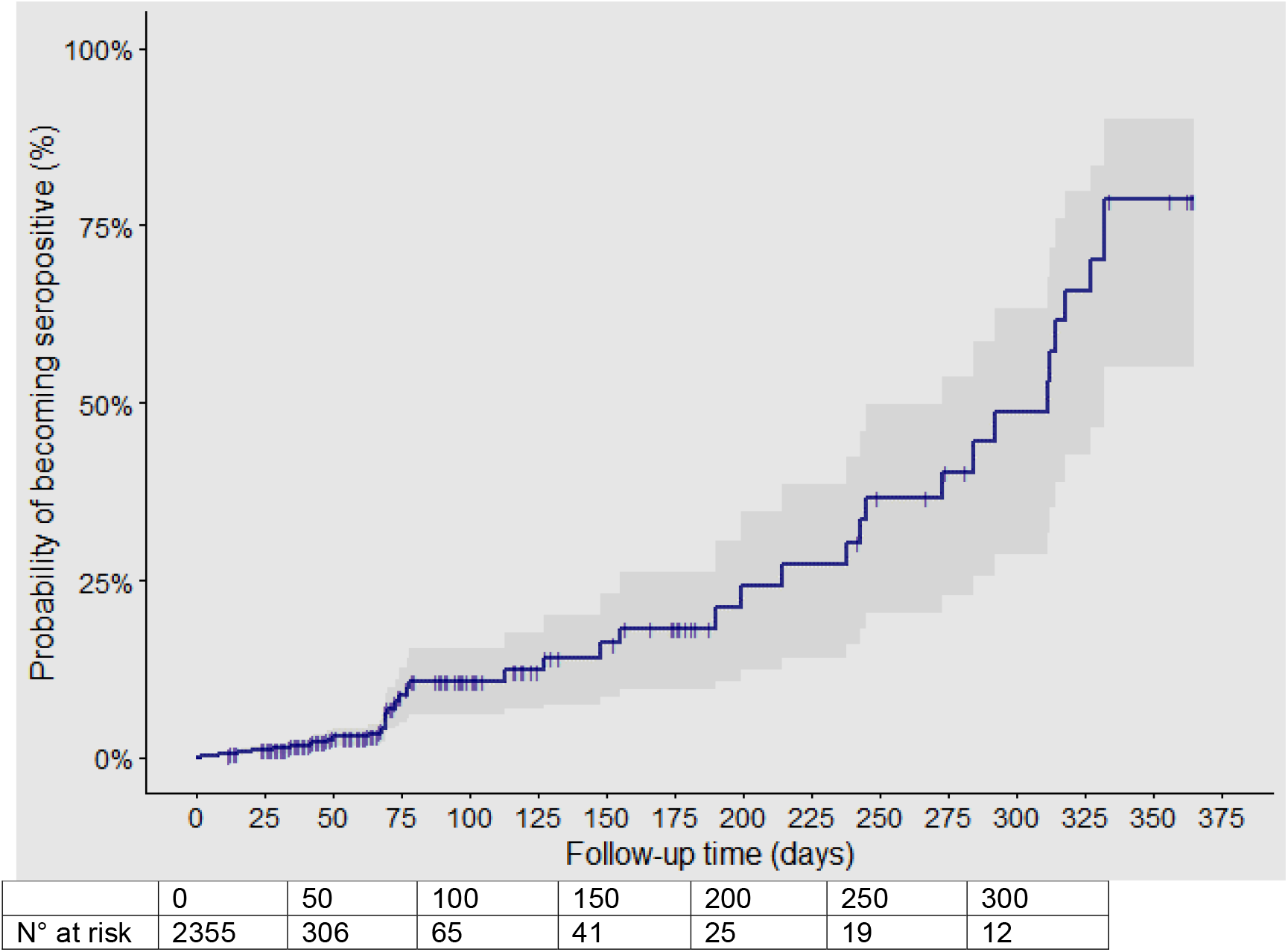
Kaplan-Meier plot^1^ of incidence of antibodies against SARS-CoV-2 among primary healthcare providers in Belgium not yet vaccinated after self-reported COVID-19 infection. ^1^ Interval censoring is taken into account by assuming that the actual event occurred somewhere between the testing timepoint of the event and the testing timepoint before

#### Among vaccinated participants

The incidence of antibodies against SARS-CoV-2 among vaccinated PHCPs in Belgium according to their self-reported history of COVID-19 infection is shown in figure 3. The incidence of antibodies is higher in PHCPs with self-reported COVID-19 infection compared to PHCPs with no self-reported COVID-19 infection both less than seven days and seven days or more after the first and the second dose, less than seven days after the third dose, but not seven days or more after the third dose.

**Figure 3.**
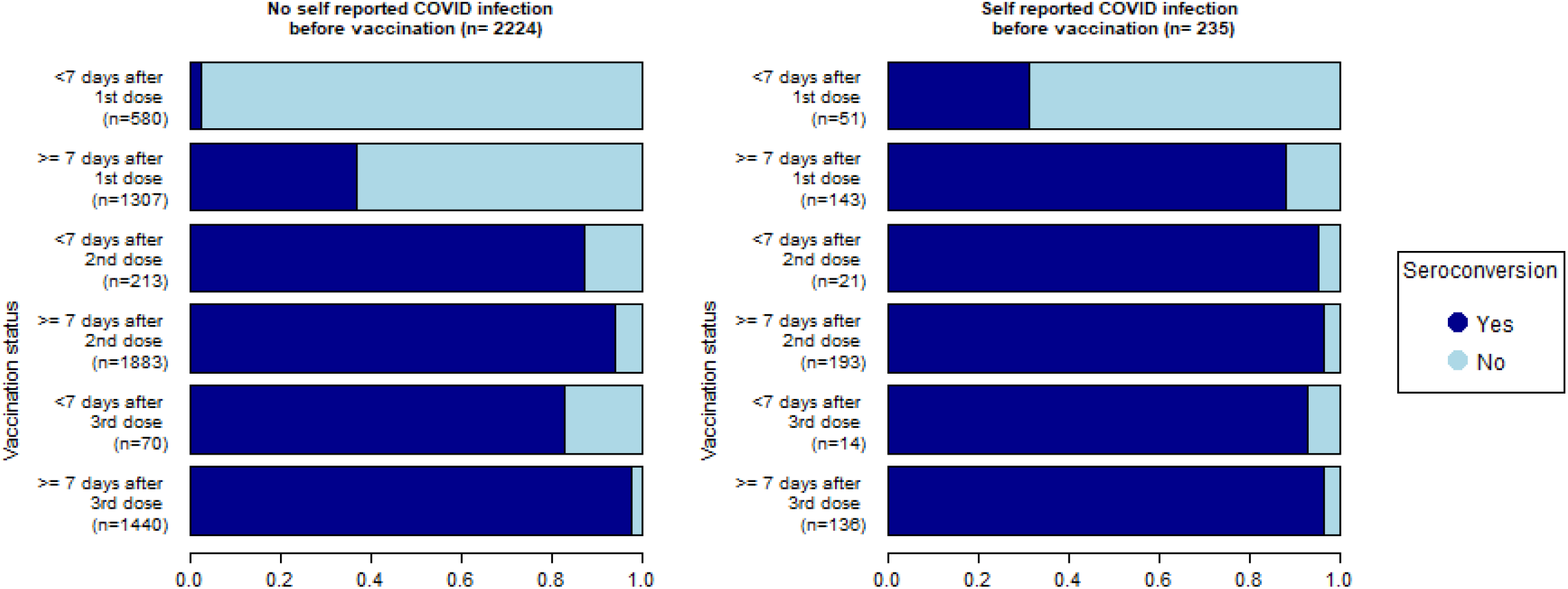
Incidence of antibodies against SARS-CoV-2 among primary healthcare providers in Belgium after vaccination according to self-reported history of COVID-19 infection

### Longevity

#### Among not (yet) vaccinated participants

The longevity of antibodies against SARS-CoV-2 among not (yet) vaccinated PHPCs in Belgium is shown in figure 4.

**Figure 4:**
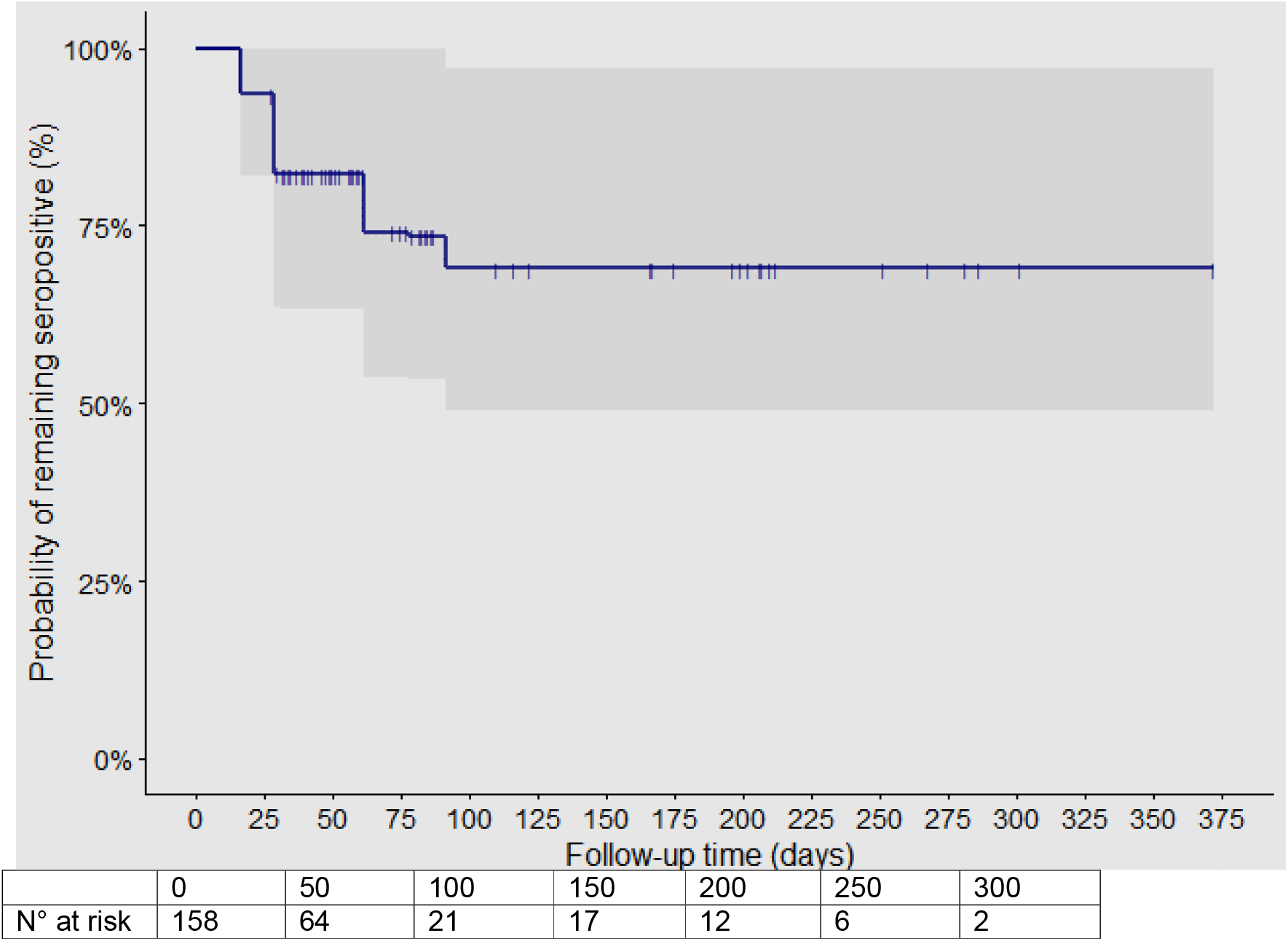
Kaplan-Meier plot^1^ of longevity of antibodies against SARS-CoV-2 among PHCPs in Belgium after self-reported history of COVID-19 infection ^1^ Interval censoring is taken into account by assuming that the actual event occurred somewhere between the testing timepoint of the event and the testing timepoint before

For T2 the positivity of antibodies against SARS-CoV-2 was estimated to be 18.54% (95%CI: 12.84-24.24; n=178)) lower compared to T1, i.e. the proportion of participants not yet vaccinated at T1 and testing positive at T1 for SARS-CoV-2 antibodies that tested negative for SARS-CoV-2 antibodies at T2. For T3 and T4 it was estimated to be 19.42% (95%CI: 11.76-27.07; n=103) and 12.50% (95%CI: 0.99 - 24.01; n=32), respectively. As of T4, the sample size of eligible participants was too small for precise estimates.

#### Among participants after full primary vaccination

The longevity of antibodies against SARS-CoV-2 among PHCPs in Belgium who have received their full primary vaccination, but not yet a booster vaccination, according to their self-reported history of COVID-19 infection is shown in figure 5. The longevity of antibodies is higher in PHCPs with self-reported COVID-19 infection compared to PHCPs without self-reported COVID-19 infection after full primary vaccination.

**Figure 5:**
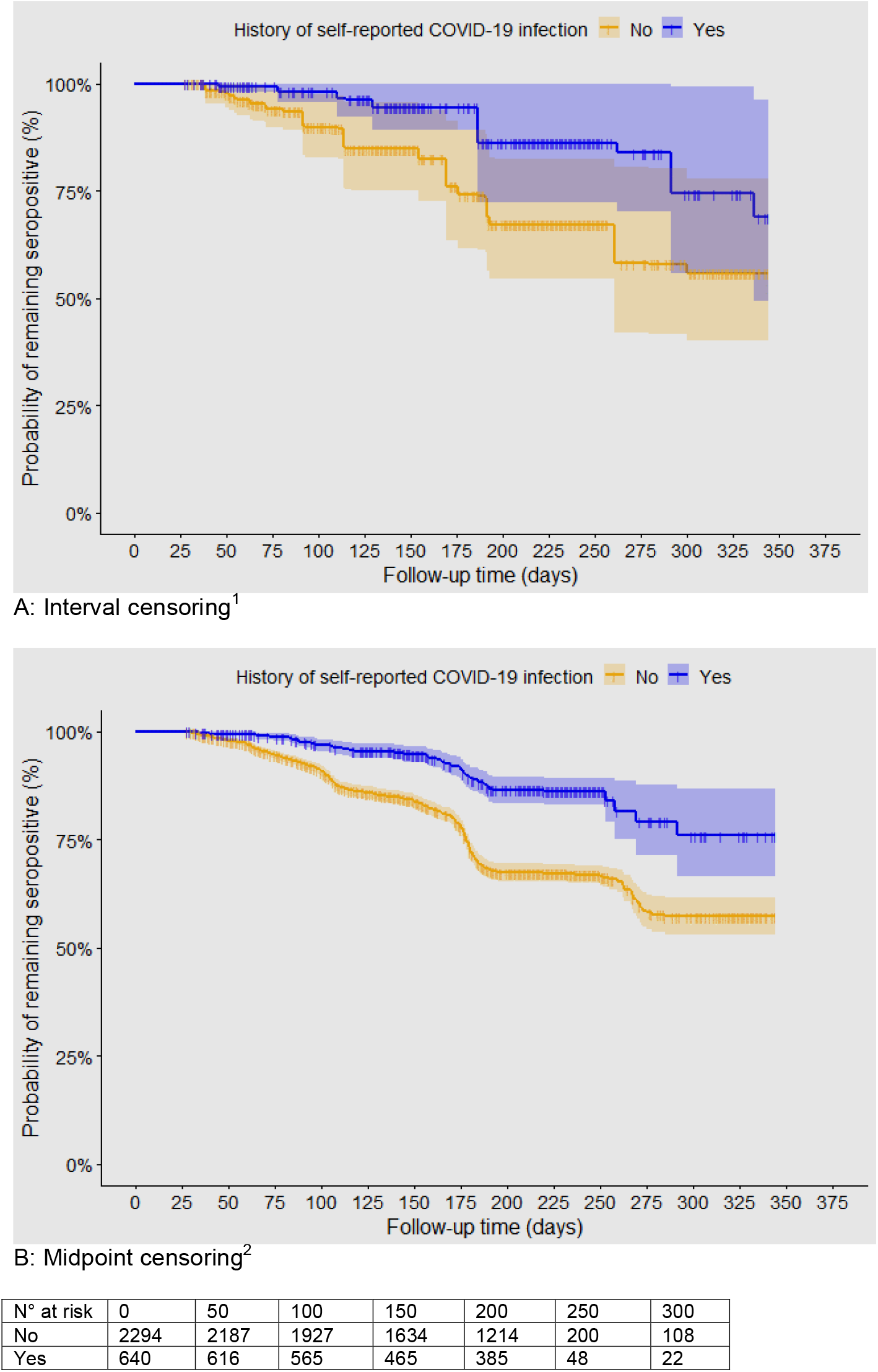
Kaplan-Meier plots of longevity of antibodies against SARS-CoV-2 among primary healthcare providers in Belgium after full primary vaccination according to self-reported history of COVID-19 infection accounting for censoring as of the booster vaccination ^1^ Assuming that the actual event occurred somewhere between the testing timepoint of the event and the testing timepoint before; ^2^ Assuming that the actual event occurred exactly between the testing timepoint of the event and the testing timepoint before.

## Discussion

The prevalence of antibodies against SARS-CoV-2 among PHCPs in Belgium was 15.1% in December 2020, i.e. before vaccination had started and right after the second Belgian COVID-19 wave that peaked beginning November 2020, and reached 93.9% in December 2021, i.e. after booster vaccination had started and after the fourth Belgian COVID-19 wave in which the Delta variant was dominant and that peaked beginning December 2021. The incidence of antibodies against SARS-CoV-2 within two weeks after COVID-19 vaccination with a first dose was higher in PHCPs with a self-reported history of COVID-19 infection compared to those with no self-reported history of infection. The longevity of antibodies was more pronounced in the former group of PHCPs than in those with no self-reported history of infection.

The seroprevalence in PHCPs before vaccination (15.1%) appeared to be lower than that among the general population (18.7%) and that among hospital health care workers (19.7%) in Belgium, in December 2020, when the Belgian healthcare system was approaching the end of the second COVID-19 wave.^15,18^ It should however be noted that the accuracy of the RST might be lower when used by many different PHCPs instead of a few trained and experienced staff (for validation) and lower than analysis of a serum sample in the lab (for seroprevalence in the general population and in hospital health care workers) using conventional lab-tests. This is suggested by the lower seroprevalence in this study for PHCPs in Flanders compared to that in an earlier prospective cohort study using dried blood spots analysed in the lab.^25^ Not finding a higher seroprevalence among PHCPs, generally concerned about being at high risk of COVID-19 infections, compared to the general population might be explained by the availability and proper usage of personal protective equipment (PPE).^25^

Most PHCPs in our study (94.49%) received a first vaccine dose in the period January – March explaining the increase in seroprevalence to 84.1% in April 2021. The monthly incidence of antibodies due to natural infection in those not yet vaccinated in the same time period was estimated to be around 4% in this study. Natural course of infection could therefore not have caused a similar rise in seroprevalence.

A gradual decrease in the prevalence of anti-SARS-CoV-2 antibodies among PHCP was observed in the following months leading to a seroprevalence of 70.2% in September 2021. In December 2021 most PHCPs (86.5% of participants in testing timepoint 8) already received a booster dose of a COVID-19 vaccine resulting in a seroprevalence of 93.1% at the end of the study. Although, also the circulation of Delta variant corona virus might have impacted this increase in seroprevalence. For example, the seroprevalence in mainly unvaccinated schoolchildren in Belgium almost doubled during the fourth covid wave (26.6% at 8 October 2021 versus 50.9% at 15 December 2021).^18, 28^ Natural infection before vaccination did seem to limit waning of antibodies after vaccination. These findings strengthen the accruing evidence base for reduced protection from infection in vaccinated, but previously uninfected participants.^29^ The clinical significance is however still to be determined. A reduction in vaccine effectiveness against infection could increase transmission to and the risk of infection among high-risk persons who consult PHCPs, some of whom may have progression to severe disease. In addition, recent studies have shown that vaccination confers more durable protection against severe outcomes of hospitalization and death than against mild symptomatic and asymptomatic infection.^30-32^ At this point studies suggest that a third or booster dose provides additional protection on top of simply reversing previous waning, but that the greatest protection from the worst clinical outcomes still remains heavily concentrated in the first two doses.^32-36^

Although studies suggest prolonged protection, it remains unclear to what extent the presence of antibodies (against the RBD) is associated with protection against new variants of the coronavirus.^36,37^ Neutralising antibody titers measured in the laboratory remain the strongest correlate of protection against symptomatic and severe illness across multiple variants.^38, 39^

This large cohort study with 12 months follow-up provided precise estimates of the prevalence and incidence of antibodies against SARS-CoV-2 among PHCPs at national and regional level. Another strength of this study is the use of RSTs. This substantially improved the timeliness of the test result availability and allowed the PHCPs to immediately check their results, which was not the case in our previous work that used dried blood spots (DBS) to assess the prevalence and incidence of antibodies against SARS-CoV-2 among PHCPs in Flanders.^23^ Consequently, the results in PHCPs in Belgium could be compared much faster to that of the general population and other population groups, e.g., health care workers in hospitals and nursing homes.

In addition, the RST used in this study allowed us to estimate the incidence and longevity of antibodies against SARS-CoV-2 both after natural infection and after vaccination. This, on the other hand, also limits seroprevalence studies like ours and others,^16^ using an RST not able to distinguish antibodies after natural infection (with new variants) from antibodies after vaccination, to assess virus circulation once the target population is highly vaccinated.

Loss to follow-up or missing data, reduced accuracy of the RST in primary care and the use of a convenience sample could also have limited the validity of the study results. However, overall retention and response of PHCPs in the study was good to excellent, we used the best available RST to avoid under- and overestimation of the presence of SARS-CoV-2 among PHCPs due to imperfect testing methods (imperfect sensitivity and specificity), and the estimates were corrected for clustering and potential geographical misrepresentation of the PHCPs.

Selection bias is possible, because the study started at the end of the second COVID-19 wave: if all the most vulnerable PHCPs had already been infected at the time of the start of this study, then the incidence among the remaining PHCPs may be lower (because better immune system, more adherent to personal protection guidelines etc.). Therefore, we explicitly asked for participation regardless of previous SARS-CoV-2 testing and test results.

In conclusion, this national study confirms results from an earlier study at regional level (Flanders only) that for the PHCPs seroprevalence and incidence during the second COVID-19 wave was similar to that of the general population suggesting that the occupational health measures implemented provided sufficient protection when managing patients. A vaccination programme including one booster increased the seroprevalence of antibodies against SARS-CoV-2 leading to a seroprevalence of 93.9% in December 2021. Between primary and booster vaccination longevity of antibodies was more pronounced in PHCPs with a history of self-reported COVID-19 infection. Therefore, continued monitoring of the seroprevalence in PHCPs after booster vaccination, with longer time intervals, could be relevant, provided that the presence of antibodies is associated with protection.

## Data Availability

All data produced in the present study are available upon reasonable request to the authors

## Authors’ contributions

The study concept and design was initiated by SC, NA, BS and ED and finalized with contributions from JYV, ADS, SH, AVdB, ID, PVD, HG. SC, NA,BS and PVN conducted registration and data collection. Analysis was performed by RB. NA prepared the first draft of the manuscript. All authors (NA, BS, RB, PVN, JYV, ADS, SH, AVdB, ID, PVD, HG, LB, ED and SC) provided edits and critiqued the manuscript for intellectual content, approved the submitted version, were involved in the interpretation of data, and agree to be accountable for all aspects of the work.

NA and BS contributed equally to this work as first author. ED and SC contributed equally to this work as last author.

## Funding statement

**‘**This work was supported by Sciensano, grant number [OZ8478]’ JYV was further supported by the National Institute for Health and Care Research (NIHR) Community Healthcare MedTech and In Vitro Diagnostics Co-operative at Oxford Health NHS Foundation Trust. The views expressed are those of the author(s) and not necessarily those of the NHS, the NIHR or the Department of Health and Social Care.

## Competing interests statement

None declared.

## Notes

### Competing Interest Statement

The authors have declared no competing interest.

### Clinical Protocols

https://www.sciensano.be/en/biblio/prevalence-and-incidence-antibodies-against-sars-cov-2-among-primary-healthcare-providers-belgium

